# Daily steps are a predictor of, but perhaps not a modifiable risk factor for Parkinson’s Disease: findings from the UK Biobank

**DOI:** 10.1101/2024.08.13.24311539

**Authors:** Aidan Acquah, Andrew Creagh, Valentin Hamy, Alaina Shreves, Charilaos Zisou, Charlie Harper, Stefan Van Duijvenboden, Chrystalina Antoniades, Derrick Bennett, David Clifton, Aiden Doherty

## Abstract

**Importance:** Higher physical activity levels have been suggested as a potential modifiable risk factor for lowering the risk of incident Parkinson’s disease (PD). This study uses objective measures of physical activity to investigate the role of reverse causation in the observed association.

**Objective:** To investigate the association between accelerometer-derived daily step count and incident PD, and to assess the impact of reverse causation on this association.

**Design:** This prospective cohort study involved a follow-up period with a median duration of 7.9 years, with participants who wore wrist-worn accelerometers for up to 7 days.

**Setting:** The study was conducted within the UK Biobank, a large, population-based cohort.

**Participants:** The analysis included 94,696 participants aged 43-78 years (56% female) from the UK Biobank who provided valid accelerometer data and did not have prevalent PD.

**Exposure:** Daily step counts were derived using machine learning models to determine the median daily step count over the monitoring period.

**Main Outcomes and Measures:** The primary outcome was incident PD, identified through hospital admission and death records. Cox proportional hazards regression models estimated hazard ratios (HR) and 95% confidence intervals (CI) for the association between daily step count and incident PD, with adjustments for various covariates and evaluation of reverse causation by splitting follow-up periods.

**Results:** During a median follow-up of 7.9 years (IQR: 7.4-8.4), 407 incident PD cases were identified. An inverse linear association was observed between daily step count and incident PD. Participants in the highest quintile of daily steps (>12,369 steps) had an HR of 0.41 (95% CI 0.31-0.54) compared to the lowest quintile (<6,276 steps; HR 1.00; 95% CI 0.84-1.19). A per 1,000 step increase was associated with an HR of 0.92 (95% CI 0.89-0.94). However, after excluding the first six years of follow-up, the association was not significant (HR 0.96, 95% CI 0.92-1.01).

**Conclusions and Relevance:** The observed association between higher daily step count and lower incident PD is likely influenced by reverse causation, suggesting changes in physical activity levels occur years before PD diagnosis. While step counts may serve as a predictor for PD, they may not represent a modifiable risk factor. Further research with extended follow-up periods is warranted to better understand this relationship and account for reverse causation.

**Key Points:** *Question:* Is there an association between accelerometer-derived daily step count and the risk of incident Parkinson’s disease, considering the potential impact of reverse causation?

*Findings:* In this prospective cohort study of 94,696 UK Biobank participants, an inverse association was found between daily step count and incident Parkinson’s disease over a median follow-up of 7.9 years. However, this association was attenuated when excluding the first six years of follow-up.

*Meaning:* While daily step counts may be associated with incident Parkinson’s disease, these observations are likely influenced by reverse causation, indicating changes in daily behaviours years before diagnosis.

## Introduction

Parkinson’s disease (PD) is the second most prevalent and most rapidly growing neurodegenerative disorder^1,2^, with an estimated prevalence of 9.4 million^3^ cases in 2020, marking a substantial increase from 5.2 million cases in 2004^4^. The prodromal phase of PD has many well-recognised motor-related symptoms, such as subtle motor dysfunction, that are believed to occur over a decade before diagnosis^5^. Recognising these early indicators could provide crucial insights for improved disease understanding and may offer the possibility of identifying modifiable risk factors for developing PD.

In existing literature, low self-reported physical activity has been associated with a higher risk of incident PD^6,7^. Despite these observations, uncertainties remain, stemming from a reliance on self-report measurements of physical activity, which are crude and less reliable, creating greater uncertainties in observed associations^8,9^. In addition, PD’s progressive and prolonged development poses challenges in distinguishing between incident and prevalent cases. This ambiguity can introduce reverse-causation bias, where the observed associations may be influenced by the change in behaviour of individuals who already have underlying PD, rather than vice-versa. One common approach to mitigate potential reverse causation is to exclude the first few years of follow-up data. In PD studies, there is inconsistency around the number of years removed across studies, ranging between four to ten years however reduced activity levels were still significantly associated with incident PD after this exclusion^6^.

Daily step counts serve as a valuable proxy for measuring physical activity levels. One of the primary advantages of using average daily steps over other physical activity metrics is their simplicity, allowing for clear communication and understanding among the general population. This ease of interpretation helps in translating research findings into effective public health messaging^10^. Existing research into the association between physical activity and incident PD have typically used metabolic equivalent of task (MET) scores to estimate physical activity^6^. However, the increasing adoption of wearable devices, capable of estimating step counts, has encouraged research on their accuracy and reliability, particularly in populations diagnosed with PD^11,12^. The translatable nature of daily step counts encourages the expansion of this research into incident PD, allowing for results to offer insight into public health recommendations for potential disease prevention.

We, therefore, aimed to investigate the association between accelerometer-measured daily step counts and incident Parkinson’s disease, and how this association changed with latter windows of follow-up.

## Methods

### Study Cohort

The UK Biobank is a prospective cohort study of 502,536 adults in the United Kingdom who were recruited between 2006 and 2010. The consented participants completed a questionnaire, interview, and provided physical measurements, as well as blood, urine, and saliva samples^13,14^. A subgroup of participants consented to wear an Axivity AX3 wrist-worn accelerometer for up to 7 days between 2013-2015^15^. All participants provided written informed consent and the study was approved by the National Information Governance Board for Health and Social Care and the National Health Service North West Multicentre Research Ethics Committee (06/MRE08/65).

### Accelerometer-derived physical activity processing

Accelerometer data was processed to derive step counts using the OxWearables “stepcount” package (version 2.1.5), a hybrid self-supervised machine learning model that was trained on ground truth free-living data and counts peaks in detected walking windows of behaviour^16^. Steps were predicted for each participant’s full period of collected data, with missing periods due to non-wear imputed by averaging the step counts in the corresponding times across valid days^16^. We then calculated the median of the imputed daily step counts, across the period of monitoring, as the primary exposure in this study.

### Ascertainment of Parkinson’s disease

The diagnosis of PD was obtained from a combination of the UK Biobank’s linked hospital data; inpatient hospital admissions data, and death data. Diagnoses were coded according to the International Classification of Diseases (version 10) coding system, with outcomes recorded as the first instance of a G20 diagnostic code^17^. This follows the approach recommended by the UK Biobank^17^, and used by other studies in studying PD in the physical activity monitoring cohort^18–20^. Incident PD however was defined at their first diagnosis of PD. Participants were censored at the first date of PD diagnosis, date of death, or at the end of their follow-up period (31 October 2022 for England, 31 August 2022 for Scotland, and 31 May 2022 for Wales), whichever came first^21^.

### Statistical Analysis

We processed the accelerometer data from 103,614 participants. We excluded participants with device calibration or data reading errors (>1% of values outside +/-8g range), inadequate wear time (<3 days, or non-wear at a particular time of day for all days of monitoring), unreasonably high average acceleration (>100 mg), and stepcount model execution failure^16,18,22,23^. We also excluded all 150 prevalent PD and 22 other Parkinsonism cases, diagnosed before accelerometer wear, and participants with missing healthcare linkages or covariate data. The final analysis included 94,696 participants (Figure 1).

**Figure 1:**
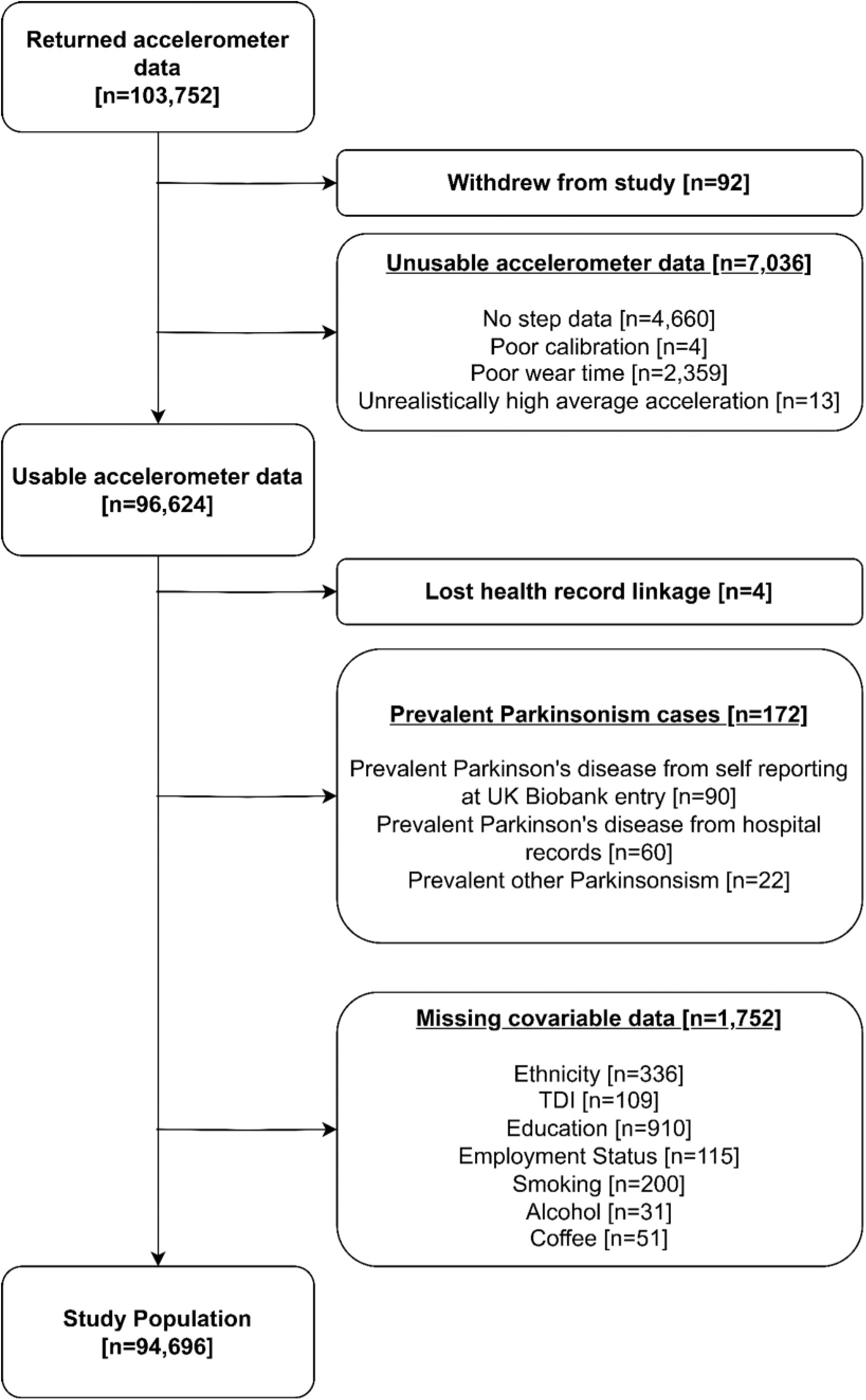
Exclusion flow diagram for analysis of UK Biobank Physical Activity Monitoring population. TDI = Townsend deprivation index.

**Figure 2:**
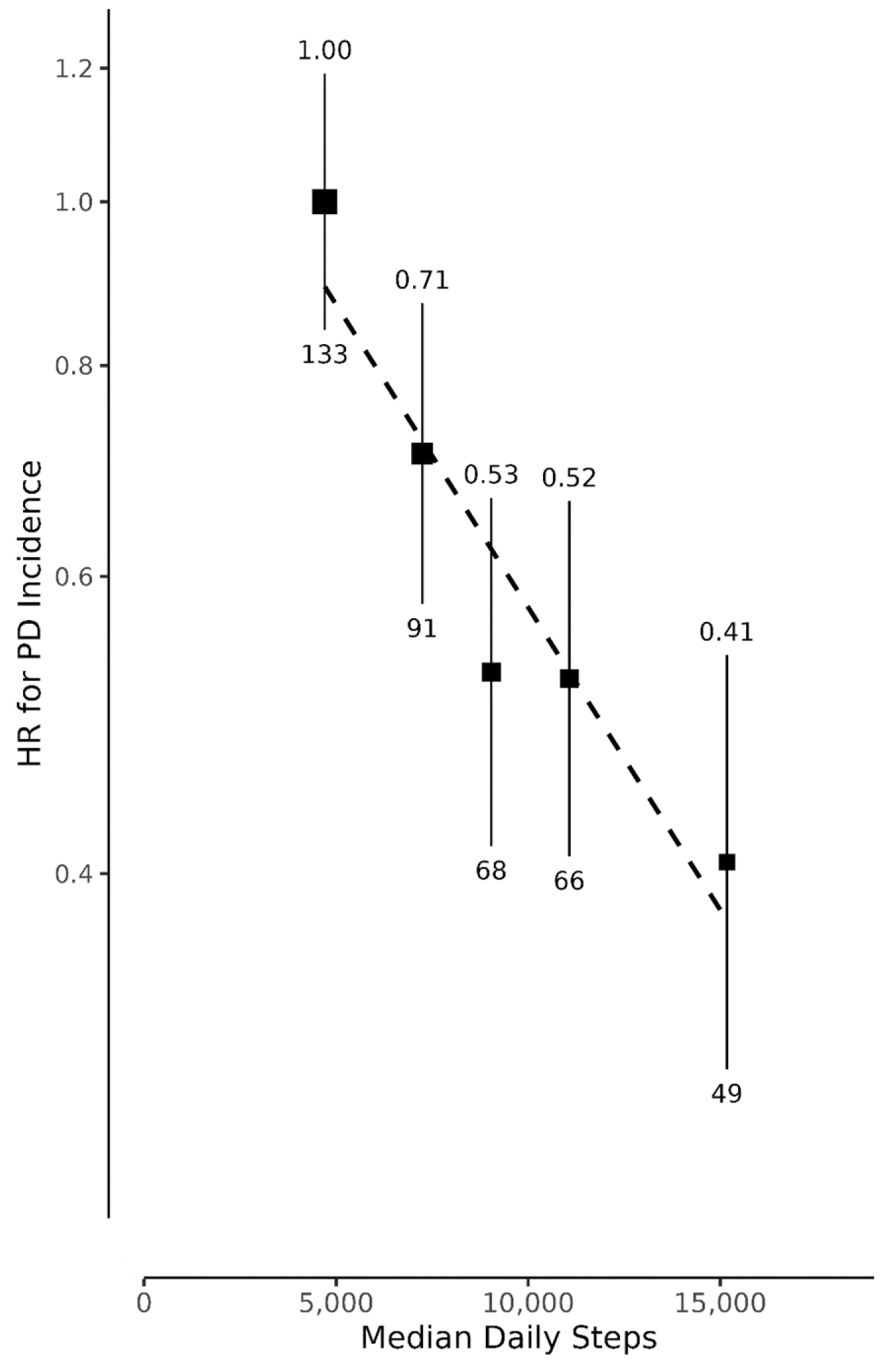
Association between quintiles of median daily step count and incident Parkinson’s disease. Hazard ratios (HR) and 95% confidence intervals were calculated using age as a timescale, adjusted for season of wear, sex, ethnicity, Townsend deprivation index, geographic region, education, employment, alcohol intake, smoking status and coffee intake. HR is above and the number of events is plotted below each data point. The HR (95% CI) per 1,000 increase in daily steps is 0.92 (0.89 – 0.94).

A Cox proportional hazard regression model^24^ was used to estimate adjusted hazard ratios (HRs) with 95% confidence intervals (CIs) for the association between daily step counts and incident PD. The association was first evaluated using quintiles of median daily step count. CIs were calculated using group-specific log HR variances to enable comparisons across all groups, presented as floating absolute risks^25^.

We then evaluated how the risk of PD was associated with each 1,000 steps per day higher. We sequentially applied the full set of adjustments to the multivariable model. The impact of these sequential adjustments were assessed using chi-squared test. Specifically, for the per-1000 median daily steps with one degree of freedom, we compared the chi-squared values before and after adjustment. A likelihood ratio test comparing quintiles of daily steps as categorical and ordinal was performed to test that the association between daily steps and incident PD did not significantly depart from linearity^26^. This allows for the reporting of hazard ratios per 1,000 steps rather than quintiles of step count. At each step of this sequential adjustment, confounding was assessed by observing the chi-squared from a likelihood ratio test of the association.

Multivariable models used age as the time scale were adjusted for season of wear (spring, summer, autumn, winter), sex (female, male), ethnicity (white, non-white), Townsend Deprivation Index (TDI, stratified by quintiles in UK population^27^), geographic region of assessment centre (London, West Midlands, Yorkshire, Northeast, Northwest, Southeast, Southwest England, Scotland, Wales), educational attainment (degree, diploma, A/AS levels, professional qualification, GCSE/O levels, no degree/qualification), employment status (employed, not employed), smoking status (never, previous, current) and alcohol consumption (never, < 3 times per week, ≥3 times per week) and coffee intake (non-drinker, ≤2 cups a day, >2 cups a day). All covariate data were provided by participants at the UK Biobank baseline assessment visit, with details provided in supplementary material. The proportional hazards assumption was tested using Schoenfeld residuals and no violations were observed.

We conducted several sensitivity analyses using the models assessing the risk for an increase in 1,000 daily steps. First, we expanded our multivariable-adjusted models by additionally adjusting for body mass index (BMI) at baseline, depression, type 2 diabetes, constipation, bladder dysfunction derived through a combination of self-report and health records, and sleep duration derived from accelerometer data. We then explored whether the observed associations differed within subpopulations of interest, including by age groups, sex, BMI (<25, 25–30, 30+ kg/m^2^)^28^ and history of depression. Additionally, we performed a sensitivity analysis examining the associations after excluding participants with any history of neurological disorder.

To address concerns regarding reverse causation bias and examine how the association may have changed over successive follow up periods, we calculated the HR within specific follow-up periods (0-2 years, 2-4 years, 4-6 years, and 6+ years). Within each follow-up period, we censored events before and/or after each interval, as seen in Floud et al. (2020)^29^.

Results were reported according to Strengthening the Reporting of Observational Studies in Epidemiology (STROBE) guidelines (Supplementary Table 3)^30^.

## Results

Among the 94,696 participants, 56% were female, 97% identified as White. Most were in the least deprived TDI quintile, compared to the general UK population^27^. Those with step counts in the highest quintile (12,369+ daily steps) were younger, had a lower TDI, and lower BMI compared to those in the lowest step count group (<6,276 daily steps).

Participants had a median follow-up time of 7.9 years (IQR: 7.4, 8.4), during which 407 incident PD cases occurred, with a median time to diagnosis of 5.2 years (IQR: 3.1, 6.8). We observed a strong and significant inverse linear association between median daily step quintiles and incident PD using data from the entire duration of follow-up. Those who walked over 12,369 steps daily had a 59% lower risk of PD (HR 0.41; 95% CI: 0.31-0.54), compared to those walking less than 6,276 steps daily (HR 1.00; 95% CI: 0.84-1.19), after multivariable adjustment. There was no evidence of departure from linearity (p=0.65).

For the multivariable model, a 1000-step increase in daily step counts was associated with an 8% lower risk of PD (HR 0.92; 95% CI 0.89-0.94). The sequential adjustment of covariates from the univariable to multivariable model showed minimal changes to the strength of the association and the chi-squared increased by 33% (shown in the supplementary material). The further adjustment of additional covariates minimally altered the observed associations. However, adjustment for sleep duration did attenuate the strength of association, with a reduction in the chi-squared by 64%, as shown in supplementary figure 4. Furthermore, we observed no significant difference across subgroups, including age groups, sex, BMI status and history of depression. After removing 9,734 individuals with prevalent neurological cases, the observed association was similar to that observed in our main study sample (HR 0.92; 95% CI 0.89-0.95).

To address potential reverse causality, we assessed how the association between step counts and PD risk may have changed over follow-up periods of <2, 2-4, 4-6 and 6+ (Figure 3). We observed the strongest association within the first two years of follow-up (HR 0.83; 95% CI 0.70-0.90), during which there were 55 incident cases. However, the association was attenuated for later windows of follow-up, trending towards the null. This is despite having higher relative numbers of cases in latter periods of follow-up, reducing uncertainty in estimates. For the follow-up period of over 6 years, the HR was no longer statistically significant, with a hazard of 0.96 (95% CI: 0.92 – 1.01).

**Figure 3:**
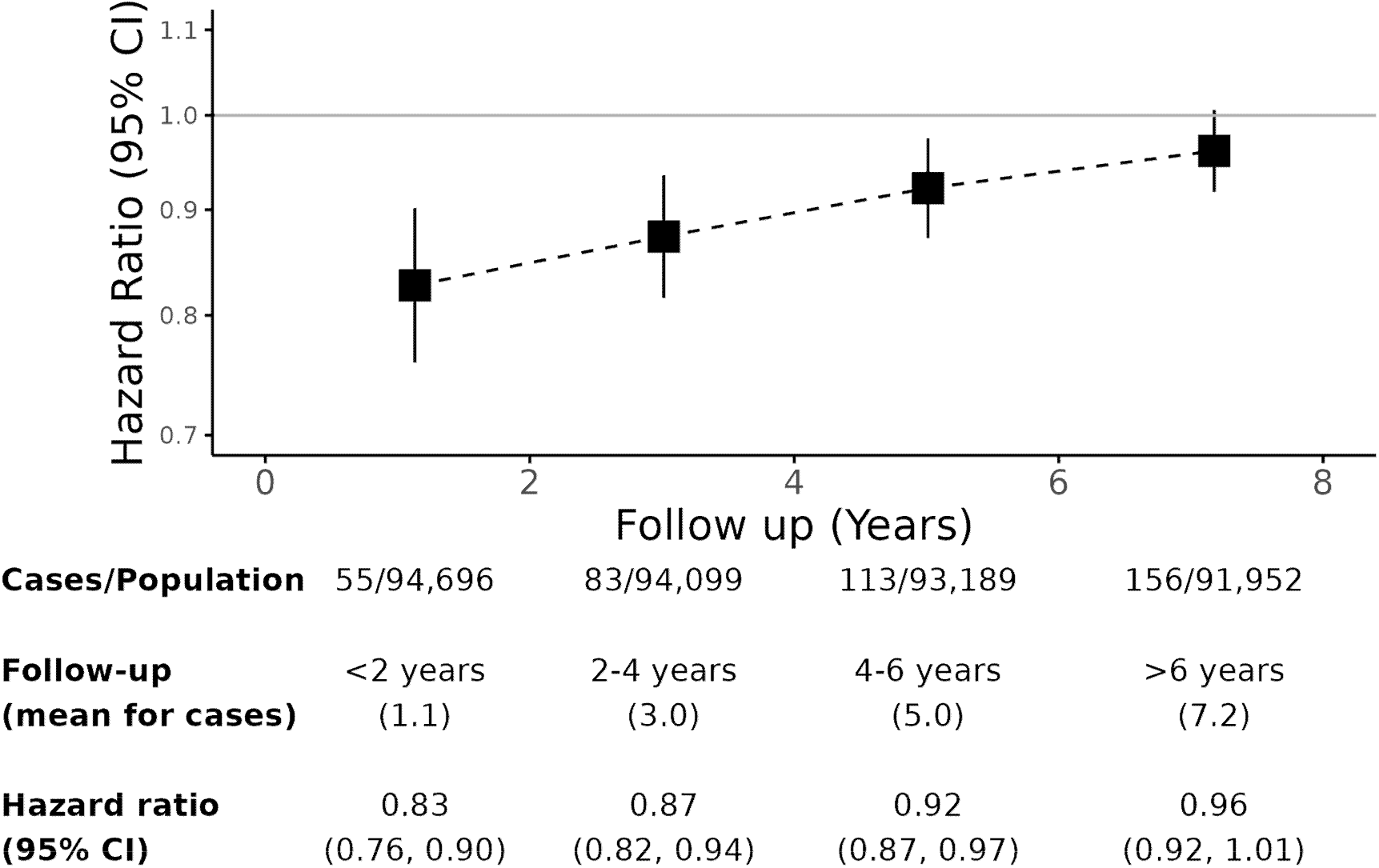
Hazard ratio for incident Parkinson’s disease by period of follow-up per 1000-step increase in daily steps. Models use age as timescale, and are adjusted for season of wear, sex, Townsend deprivation index, geographic region of assessment centre, educational attainment, employment, smoking, alcohol and coffee consumption. CI=confidence interval.

## Discussion

Higher accelerometer-derived physical activity levels, indicated by median daily step counts, were associated with a lower risk of incident Parkinson’s disease (PD) when using data from all periods of follow-up. This association remained statistically significant after multivariable adjustment, investigating potential factors, and among subpopulations of interest. However, this association was null when considering later periods of follow-up, indicating that reverse causation bias is likely. Overall, these findings do not provide evidence that daily step counts may be a modifiable risk factor for PD.

Our findings are in contrast to some previous literature in the field that supports an association between physical activity levels and incident PD, even after the consideration of reverse causation^6,19,20,31^. In a meta-analysis of this association with a median (range) follow-up of 12 (6.1-22.0) years, the relative risk for incident PD comparing the highest and lowest categories of total physical activity was 0.79 (95% CI 0.68-0.91), and remained significant when the first four to ten years of follow-up was removed from the constituent studies (0.76; 95% CI 0.64-0.91)^6^. In our study, removing years of follow-up reduced the strength of the observed association. Similar attenuations between physical activity and diseases of aging have been reported in other studies. Our findings are more consistent with results from a study on physical activity and dementia that was conducted in the Whitehall Study and Million Women Study^29,32^.

These results however support existing literature in finding lower physical activity levels to be a strong predictor for late-stage prodromal PD/early-stage clinical PD among those who already have a PD diagnosis^31,33^. One study reported that this reduction in activity may occur up to 4 years before a clinical diagnosis^31^. A separate study assessing minutes of daily physical activity^33^ found a reduction of 29% in comparing clinic PD patients to controls, in minutes of daily physical activity. There are several hypotheses for these activity changes, including changes in overall disability, poorer walking performance, and fear of falls/anxiety/depression^33^. It should be noted, however, that this reduction in activity is not fully explained by known factors, with this study encouraging further research into explaining this behaviour^33^.

Our study has several strengths. First, we used objective measures of activity derived from accelerometer data, which offers greater reliability in the measurement of this exposure compared to questionnaire-based studies. Second, the use of step count as a measure of daily physical activity offers the opportunity for better communication as step counts are easily translatable for public health messaging. Next, this work has built on previous studies investigating PD using the UK Biobank physical activity monitoring study^18–20^, however with longer follow-up, while also addressing some of these studies’ limitations. Of these limitations, we explored how the association may change over follow-up periods to better describe the potential impact of reverse causation.

There are also some limitations to our study. Firstly, this analysis was purely observational; hence we cannot exclude the possibility of unmeasured or residual confounding in the observed associations, despite considering a comprehensive set of confounders. Secondly, the UK Biobank provides limited health data linkage for participants, which we use to identify participants with PD. We use the approach recommended by UK Biobank to ascertain PD, combining hospital inpatient, death register and self-reported data^17^. However, this does not provide information regarding the severity or subtype of PD, which may offer further insights into this association. Furthermore, the use of primary care data could bolster the PD cases, but this data is currently limited to just 45% of participants and only up to 2017. Lastly, this study is limited by the number of cases observed within period of follow-up. With 407 incident cases of PD from all periods of follow-up, and just 156 diagnosed at least 6 years after participating in the study, we are somewhat limited in power to observe small effects of increased activity. We are also limited by a maximum follow-up time of 9.4 years. The PD prodromal window is typically reported to be at least 10 years prior to a clinical diagnosis^5^. Future studies in this population can benefit from increase case numbers from a longer follow-up period, while also giving further insights into this association, beyond the prodromal window of PD.

## Conclusion

Results from this prospective study add to the literature on physical activity and incident PD, suggesting that higher daily step counts were associated with lower risk of incident PD, but only for short follow-up periods.

Therefore, the observed association, may be explained by reverse causation bias. This work supports the continued labelling of low physical activity as a marker for Parkinson’s disease, rather than as a modifiable risk factor leading to Parkinson’s disease.

## Data Availability

All data produced are available online at

https://www.ukbiobank.ac.uk/enable-your-research/apply-for-access

## Acknowledgements

We thank Prof. Sarah Floud and Tonima Trisa for their guidance in the handling of reverse causation in this association.

## Appendix

**Table 1:**
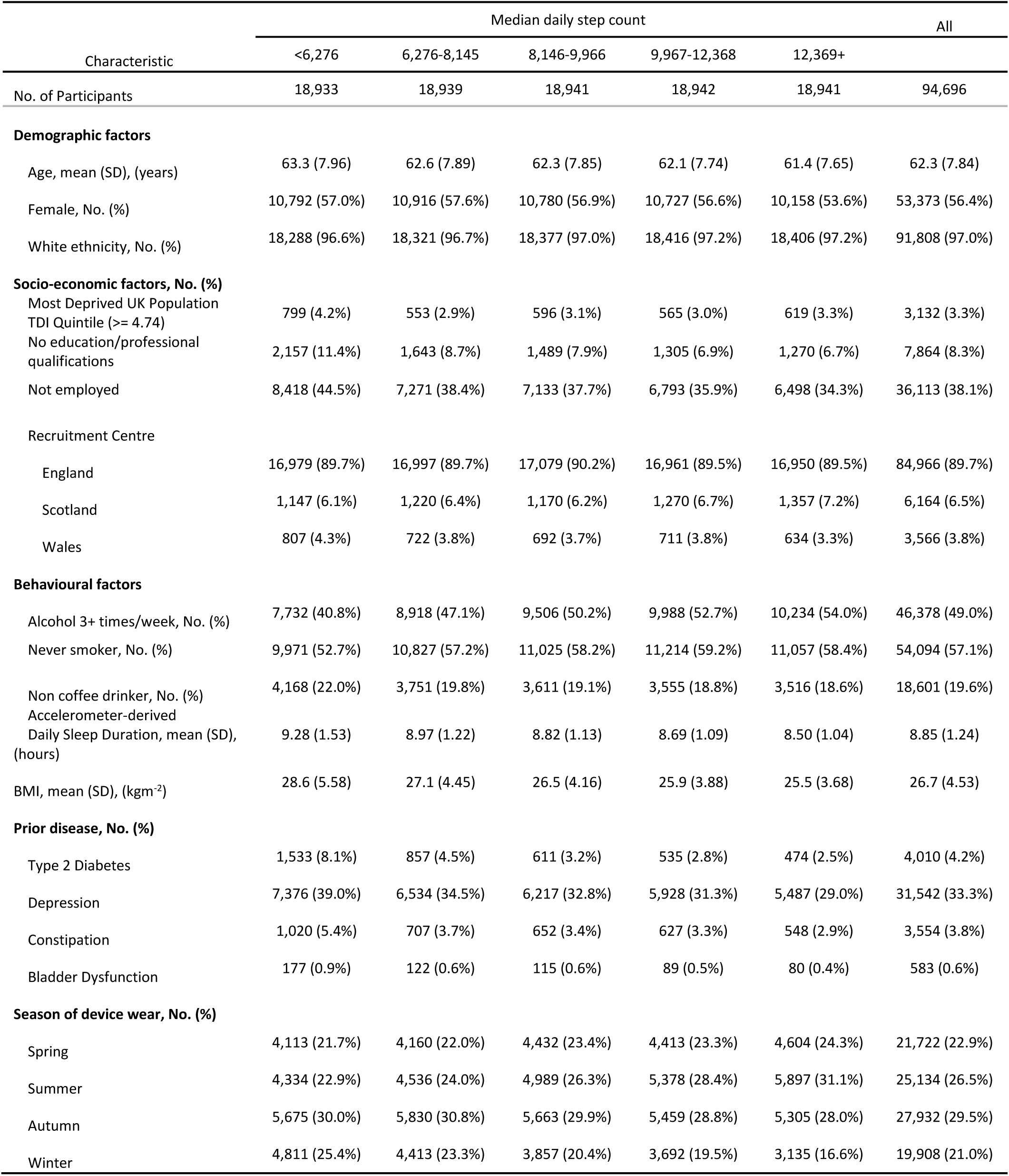
Count by Demographic Characteristics in the UK Biobank.

