## Supplementary material for "Daily steps are a predictor of, but perhaps not a modifiable risk factor for Parkinson’s Disease: findings from the UK Biobank"

Supplementary materials

**Tables**

*Supplementary Table 1: Description of datasets used in this analysis.*

| Characteristic | OxWalk study | UK Biobank Physical Activity Cohort |
|---|---|---|
| No. of participants | 39 | ~ 100,000 |
| Sensor | Axivity AX3 | Axivity AX3 |
| Raw sampling rate | 100Hz | 100Hz |
| Body location | Dominant wrist | Dominant wrist |
| Activity protocol | Unscripted free-living | Unscripted free-living |
| Measurement window, per participant | 1 hour | 7 days |
| Ground truth capture | Waist worn, foot facing, video camera | None |
| Participant Age | 38.5 (SD 14.0)<br>Range: 19 - 81 | 62.4 (SD 7.8)<br>Range: 43 - 79 |
| Participant Sex | 19 Female, 20 Male | ~56,000 Female,<br>~44,000 Male |

*Supplementary Table 2: Definition of variables from the UK Biobank data*

| Characteristic | Source | Notes | UK Biobank field | Coding Notes |
|---|---|---|---|---|
| **OUTCOME** | | | | |
| **Age at first PD event** | Hospital episode statistics (HES) records | First ICD-10 code (G20) in HES data. | Derived from 2000 | |
| **Age loss-to-follow up** | Death Registry | | Derived from 100093 | |
| **EXPOSURE** | | | | |
| **Median Daily Step count** | Accelerometry | Derived using the "stepcount" tool, version 2.1.5.<br>Details can be found at:<br>https://github.com/OxWearables/stepcount | | Reported either as divided into tertiles:<br><7,560, 7,560-10,646, 10,646+<br>Or as per-1000 daily steps |
| **EXCLUSION VARIABLES** | | | | |
| **Prior Parkinsonism** | HES records | PD-related ICD-10 codes in hospital records before accelerometry wear date:<br>G20-26, G35, G47 | Derived from 2000 | |
| | Baseline | PD-related self-reported disease | Derived from 20002 | |
| | Algorithmically derived first instance of PD (G20) | Date G20 first reported (Parkinson's disease) prior to the accelerometry wear date | Derived from 131022 | |
| **ADJUSTMENT VARIABLES** | | | | |
| **Age** | Baseline | Attained age was the underlying timescale in survival analyses; participants entered the study at the end of accelerometer wear. | Derived from 90011, 34, 52 | |

| | | | | |
|---|---|---|---|---|
| **Sex** | Baseline | | 31 | Female, Male |
| **Ethnicity** | Baseline | | Derived from 21000 | White, non-white |
| **Townsend Deprivation Index** | Baseline | Townsend Deprivation Index of address at time of UKB baseline assessment.<br>UK population quintiles: linked here | Derived from 22189 | Quintiles of UK Population |
| **UK recruitment centre geographic region** | Baseline | | Derived from 54 | London, West Midlands, Yorkshire, northeast, northwest, southeast, southwest England, Scotland, Wales |
| **Education/Qualifications** | Baseline | Selected as highest educational achievement. Labels are ordered by highest to lowest value.<br>ISCED<br>UK Biobank Paper | Derived from 6138 | College/University degree, NVQ/HND/HNC or equivalent, Other professional qualification, A levels/AS levels or equivalent, GCSEs/CSEs/O levels or equivalent, No qualification |
| **Employment status** | Baseline | | Derived from 6142, 20119 | Employed, Not employed |
| **Smoking status** | Baseline | | Derived from 20116 | Never, previous, current |
| **Alcohol consumption** | Baseline | | Derived from 1558 | Never, <3 times per week, ≥3 times per week |
| **Coffee intake** | Baseline | | Derived from 1498 | Non-drinker, Drinker - up to 2 cups, Drinker - more than 2 cups |
| **ADJUSTMENT VARIABLES – SENSITIVITY ANALYSIS** | | | | |
| **Body mass index** | Baseline | | Derived from 21001 | |
| **Average sleep duration** | Accelerometry | | Derived from 40046 | |
| **Type 2 Diabetes** | HES records | ICD-10 code (E11) in HES data prior to end of accelerometer wear. | Derived from 2000 | |
| | Baseline | Type 2 Diabetes self-reported disease at baseline | Derived from 20002, 2443 | |

| | | | |
|---|---|---|---|
| **Depression** | HES records | ICD-10 code (F32, 33) in HES data prior to end of accelerometer wear. | Derived from 2000 |
| | Baseline | Depression self-reported disease at baseline | Derived from 20002, 2090, 2100 |
| **Constipation** | HES records | ICD10 code (K59.0) in HES data prior to accelerometer wear | Derived from 2000 |
| | Baseline | Use of laxatives in self-reported medication use at baseline | Derived from 6154 |
| **Bladder dysfunction** | HES records | ICD10 code (N31) in HES data prior to accelerometer wear | Derived from 2000 |
| | Baseline | Incontinence self-reported disease at baseline | Derived from 20002 |

| | | | |
|---|---|---|---|
| **Neurological conditions** | HES records | Any of the following ICD10 codes in HES data prior to accelerometer wear:<br>A80, A80.0, A80.1, A80.2, A80.3, A80.4, A80.8, A80.9, D32, D36, E10.4, E11.4, E12.4, E13.4, E14.4, F00, F00.0, F00.1, F00.2, F00.9, F01, F01.0, F01.1, F01.2, F01.3, F01.8, F01.9, F02, F02.0, F02.1, F02.2, F02.3, F02.4, F02.8, F03, F05, F05.1, G00, G01, G02, G03, G04, G06, G07, G12, G25.0, G30, G30.0, G30.1, G30.8, G30.9, G31, G31.1, G32, G35, G36, G37, G40, G43, G44, G50, G50.0, G50.1, G50.8, G50.9, G51, G51.0, G52, G53, G57, G60, G61, G61.0, G62, G70.0, G72, G80, G81, G82, G82.2, G82.3, G82.4, G83, G83.0, G83.1, G83.2, G83.3, G83.4, G83.8, G83.9, G84, G90, G91, G92, G93, G93.0, G93.1, G93.2, G93.3, G93.4, G93.5, G93.6, G93.7, G93.8, G93.9, G94, G95, G96, G96.0, G96.1, G96.8, G96.9, G97, G98, G99, H53, H81, H81.0, H81.1, H81.2, H81.3, H81.4, H81.8, H81.9, H83, H90, H91, H91.0, H91.1, H91.2, H91.3, H91.8, H91.9, H93, H93.0, H93.1, H93.2, H93.3, H93.8, H93.9, I67, I67.0, I67.1, I67.2, I67.3, I67.4, I67.5, I67.6, I67.7, I67.8, I67.9, K56, M79, M79.0, M79.1, M79.2, M79.3, M79.4, M79.6, M79.7, M79.8, M79.9, N08, N08.0, | Derived from 2000 |

| | | |
|---|---|---|
| | | N08.8, N08.9, N13, N13.0, N13.1, N13.2, N13.3, N13.4, N13.5, N13.6, N13.7, N13.8, N13.9, Q05, R29.0, R41.81, S0[0-9], S14, S14.0, S24, S24.0, S24.1, S34, S34.0,<br>S34.1, S34.3, S44, S54, S64, S74, S84, S94, T04, T06, T07, T14, T24, T84, T90, T90.3, T90.4, T90.5, T90.8, T90.9, T91, T91.3 |

| | | | | |
|---|---|---|---|---|
| | Baseline | Any of the following self-reported diseases at baseline:<br>infection of nervous system, brain abscess/intracranial abscess, encephalitis, meningitis, spinal abscess, cranial nerve problem/palsy, bell's palsy/facial nerve palsy, trigemminal neuralgia, spinal cord disorder, paraplegia, spina bifida, peripheral nerve disorder, peripheral neuropathy, acute infective polyneuritis/guillain-barre syndrome, trapped nerve/compressed nerve, diabetic neuropathy/ulcers, chronic/degenerative neurological problem, motor neurone disease, myasthenia gravis, multiple sclerosis, parkinsons, dementia, alzheimers, cognitive impairment, other demyelinating disease, epilepsy, migraine, cerebral palsy, other neurological problem, headaches, benign / essential tremor, polio / poliomyelitis, meningioma / benign meningeal tumour, benign neuroma, neurological injury/trauma, head injury, spinal injury, peripheral nerve injury | Derived from 20002 | |
| **DESCRIPTIVE VARIABLES – DESCRIPTIVE TABLES** | | | | |
| **Wear season** | | | Derived from 90001 | Winter, Spring, Summer, Autumn |

*Supplementary Table 3: Strengthening the Reporting of Observational Studies in Epidemiology (STROBE) guidelines checklist for reporting of findings with this work. Pages with the S prefix correspond to supplementary material, otherwise refer to the main manuscript.*

| **Section** | **Subsection** | **Summary** | **Code** | **Pages** |
|---|---|---|---|---|
| Title and Abstract | Title and Abstract | Indicate study's design in title/abstract | 1a | 1-3 |
| Title and Abstract | Title and Abstract | Provide informative summary of study | 1b | 2-3 |
| Introduction | Background/rationale | Explain scientific background and rationale | 2 | 4 |
| Introduction | Objectives | State specific objectives and hypotheses | 3 | 4 |
| Methods | Study design | Present key elements of study design | 4 | 4-6 |
| Methods | Setting | Describe setting, locations, and relevant dates | 5 | 4 |
| Methods | Participants | Give eligibility criteria and selection methods | 6a | 4-5 |
| Methods | Participants | Provide matching criteria for matched studies | 6b | N/A |
| Methods | Variables | Define outcomes, exposures, confounders, and effect modifiers | 7 | 5-6 |
| Methods | Data sources measurement | Give sources and methods of data assessment | 8 | 5-6 |
| Methods | Bias | Describe efforts to address bias | 9 | 6 |
| Methods | Study size | Explain how study size was determined | 10 | N/A |
| Methods | Quantitative variables | Explain handling of quantitative variables | 11 | 5 |
| Methods | Statistical methods | Describe all statistical methods used | 12a | 5 |
| Methods | Statistical methods | Describe methods for examining subgroups and interactions | 12b | 6 |
| Methods | Statistical methods | Explain how missing data were addressed | 12c | 5 |

| | | | | |
|---|---|---|---|---|
| Methods | Statistical methods | Explain how loss to follow-up was addressed | 12d | 6 |
| Methods | Statistical methods | Describe any sensitivity analyses | 12e | 6 |
| Results | Participants | Report numbers at each stage of the study | 13a | 6 |
| Results | Participants | Give reasons for non-participation | 13b | N/A |
| Results | Participants | Consider use of a flow diagram | 13c | 16 |
| Results | Descriptive data | Provide characteristics of study participants | 14a | 6-7 |
| Results | Descriptive data | Indicate number of participants with missing data | 14b | 16 |
| Results | Descriptive data | Summarise follow-up time | 14c | 6 |
| Results | Outcome data | Report numbers of outcome events or summary measures | 15 | 6 |
| Results | Main results | Give unadjusted and adjusted estimates with precision | 16a | 7-8, 17-18, S11 |
| Results | Main results | Report category boundaries when continuous variables were categorized | 16b | S3-7 |
| Results | Main results | Translate estimates of relative risk into absolute risk if relevant | 16c | N/A |
| Results | Other analyses | Report other analyses done | 17 | 7-8 |
| Discussion | Key results | Summarise key results with reference to objectives | 18 | 8 |
| Discussion | Limitations | Discuss the limitations of the study | 19 | 9 |
| Discussion | Interpretation | Provide overall interpretation of results | 20 | 8-9 |
| Discussion | Generalisability | Discuss the generalisability of the study results | 21 | 9 |
| Other information | Funding | Give the source of funding and role of funders | 22 | 3 |

**Figures**

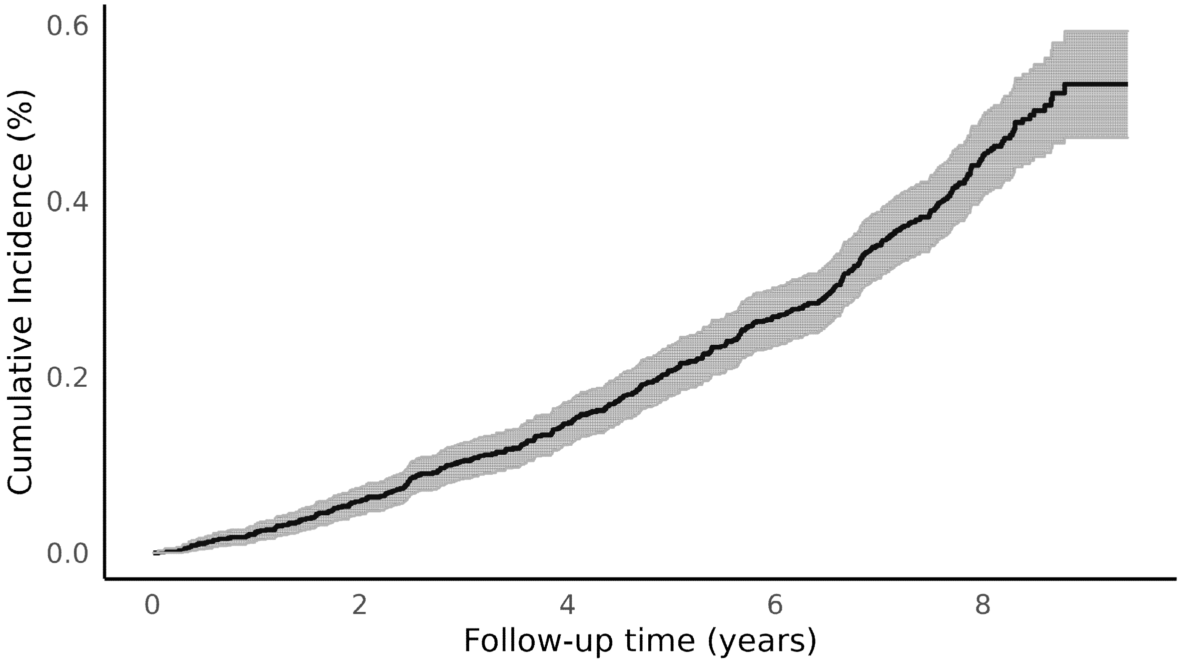

*Supplementary Figure 1: Kaplan-Meier curve for cumulative incidence of Parkinson's disease in the UK Biobank physical activity monitoring study population.*

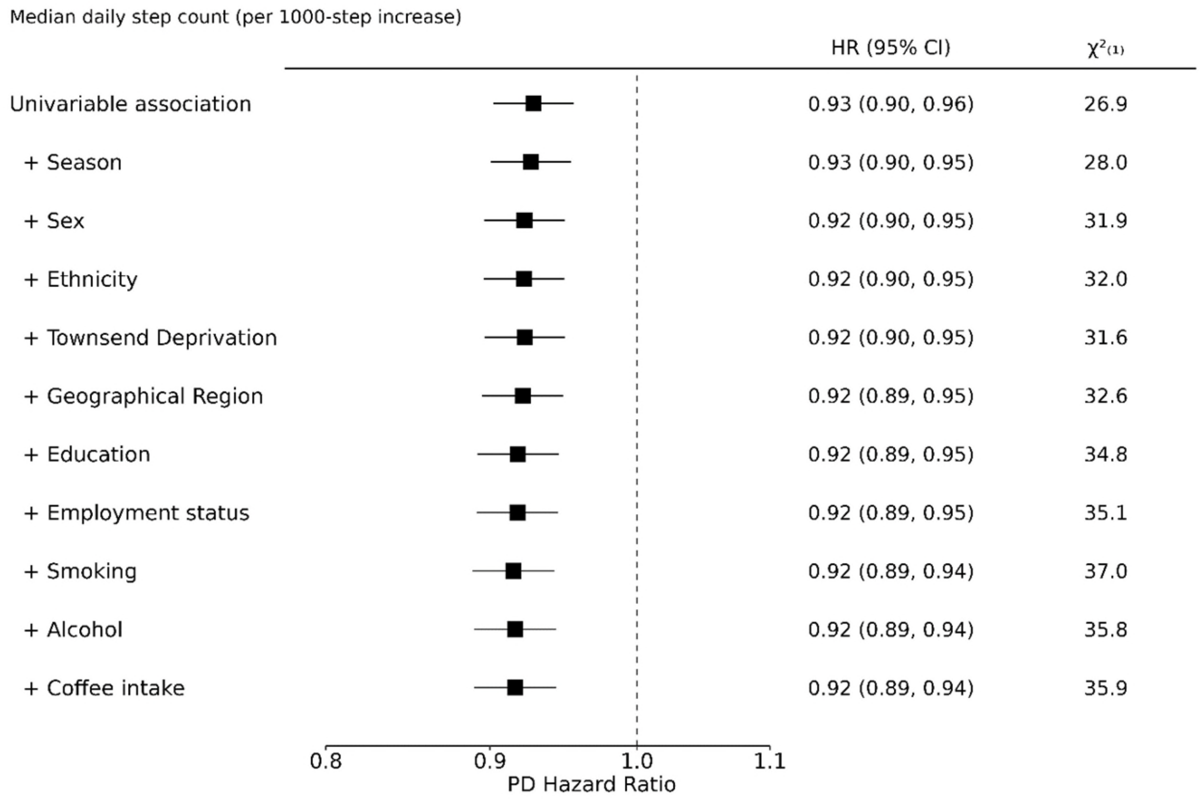

*Supplementary Figure 2: The sequential adjustment of the association between a per 1000-step increase in imputed median daily step count and incident Parkinson's disease with age as timescale using all periods of follow-up. HR=Hazard ratio, CI=Confidence interval,* $\chi^2_{(1)}$*=chi squared using likelihood ratio test for one degree of freedom.*

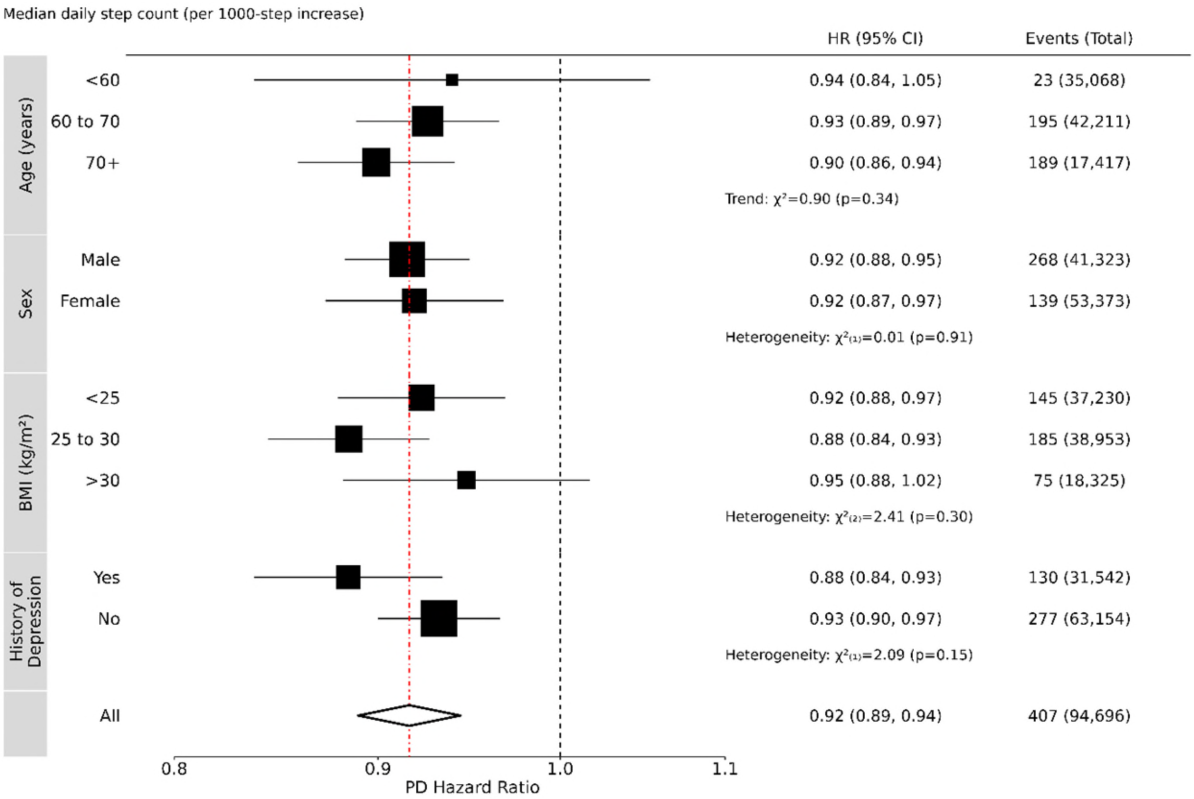

*Supplementary Figure 3: The association between a per 1000-step increase in imputed median daily step count and incident Parkinson's disease with age as timescale using all periods of follow-up within various subpopulations. Models are adjusted for season, sex (except for sex-based sub-populations), ethnicity, Townsend deprivation, geographic region, education, employment status, smoking, alcohol and coffee intake. BMI=Body Mass Index, HR=Hazard ratio, CI=Confidence interval.*

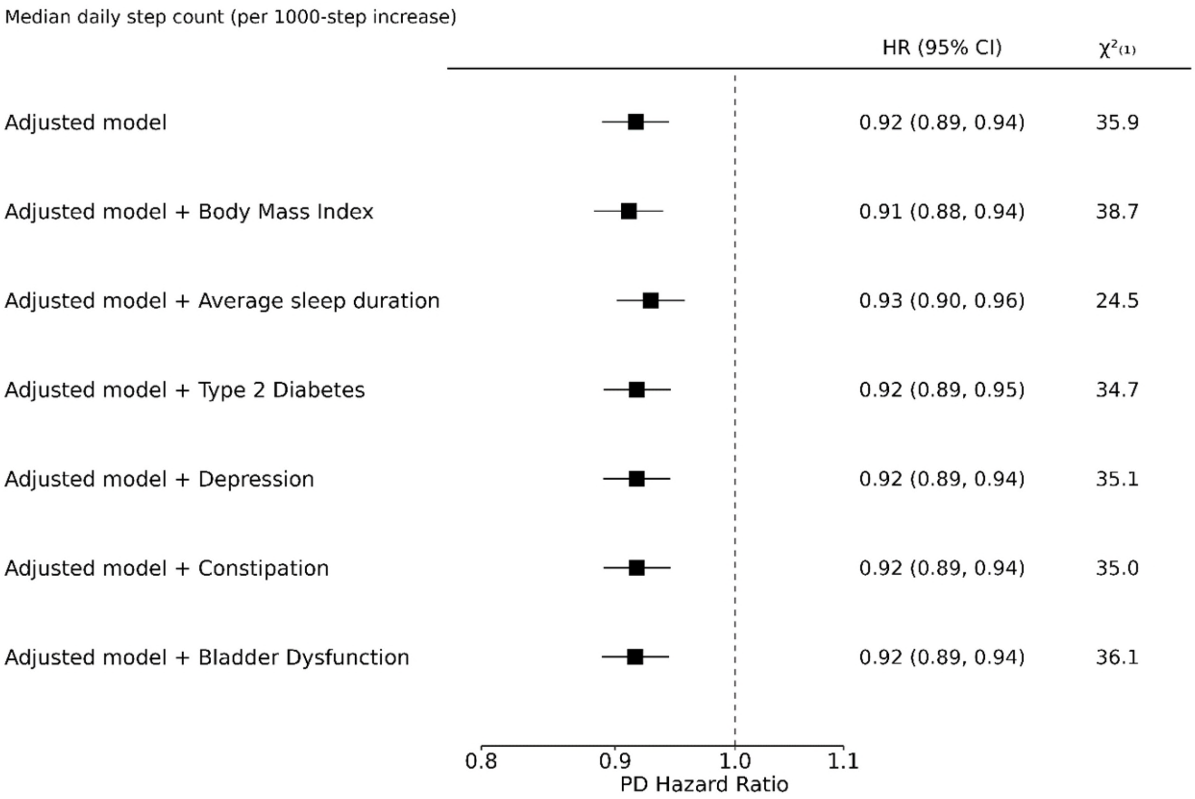

*Supplementary Figure 4: The association between a per 1000-step increase in imputed median daily step count and incident Parkinson's disease with age as timescale using all periods of follow-up after adjustment for potential confounding factors. The adjusted model is adjusted for season, sex, ethnicity, Townsend deprivation, geographic region, education, employment status, smoking, alcohol and coffee intake. HR=Hazard ratio, CI=Confidence interval,* $\chi^2_{(1)}$*=chi squared using likelihood ratio test for one degree of freedom.*
